# Human health effects of recycling and reusing plastic packaging in the food system: a systematic review and meta-analysis of life cycle assessments

**DOI:** 10.1101/2022.04.22.22274074

**Authors:** Megan Deeney, Rosemary Green, Xiaoyu Yan, Claire Dooley, Joe Yates, Heike B Rolker, Suneetha Kadiyala

**Affiliations:** London School of Hygiene & Tropical Medicine; London, United Kingdom; University of Exeter; Exeter, United Kingdom; University of Bristol; Bristol, United Kingdom; Rothamsted Research, North Wyke, United Kingdom

## Abstract

Circular strategies, including recycling and reuse of food packaging, are critical responses to the plastic pollution crisis and could provide co-benefits and trade-offs for human health. Our meta-analysis of life cycle assessment (LCA) data quantifies possible health effects using Disability-Adjusted Life Years (DALYs) mediated by climate change, ozone, air pollution, toxicity, and water scarcity. We found strong evidence for reduced health risks with both a higher percentage of recycled content and a greater end-of-life recycling rate, resulting in around a day of healthy life saved per tonne of plastic packaging recycled. On average, reusable packaging reduced the health impacts associated with single use plastics after 30 uses, which is unlikely reflected in current consumer behaviour. Data from low- and middle-income countries, and greater use of health indicators in LCA, are urgently needed. LCA is a unique tool that could be optimised for interdisciplinary public health research on circular economies.

**Teaser:** Life cycle assessment meta-analysis shows recycling and reusing plastic food packaging could provide human health co-benefits, and some risks.

## Introduction

By 2050, the plastics sector could be responsible for 20% of oil consumption and 15% of the annual global carbon budget (*1*), with single use packaging driving a cumulative 12 billion metric tonnes of plastic landfill and pollution (*2*). Food and beverage packaging may account for 65% of the plastic packaging reaching consumers (*3*) and is a central focus of circular economy initiatives promoting recycling and reuse. Whilst reducing environmental burdens, these strategies may also present risks and opportunities for human health(*4*).

Plastic offers an economic, versatile, and hygienic packaging solution for longer lasting food and drink. Since mass production of plastic began in the 1950s, global consumption has increased exponentially, outstripping that of any other manufactured material (*2*). Over 90% of plastic is still derived from finite virgin fossil feedstocks and most plastic packaging is discarded in the same year that it is produced (*1, 2*). Global average recycling rates remain low at 18% of non-fibre plastic and the rest is incinerated, landfilled, or accumulates in the environment(*2*). Plastic can take hundreds of years to degrade(*5*), its durability a gross mismatch with its brief, single use by consumers. This predominantly linear economy of plastic packaging is a resource-intensive and highly wasteful ‘take-make-dispose’ model of material management, resulting in unsustainable environmental damage and subsequent human health risks.

The Sustainable Development Goals (SDGs) put responsible management of resources and waste reduction on the global agenda, highlighting recycling and reuse as key alternatives to the linear economy (*6*). International targets for increasing recycling and reuse have been rapidly adopted in response to the plastic pollution crisis. The New Plastics Economy Global Commitment signatories, representing 20% of the plastic packaging industry and 20 government bodies in five continents (*7*), have committed to fully decoupling plastic production from finite resources and to recycling, reusing or composting 100% of packaging (*8*). The European Strategy for Plastics in a Circular Economy states that by 2030 all plastic packaging entering the European market should be either recyclable or reusable(*9*). Countries across Africa have trailblazed single use plastic bans (*10, 11*); the African Circular Economy Alliance calls for legislative changes allowing recycled plastic to be used in food grade packaging and more efficient plastic recycling trade between countries (*12*). The stage is set for a global break from single use plastic packaging, with increasing recycling and reuse around the world, though significant action across sectors will be required to realise these ambitions.

It is important that any trade-offs and co-benefits associated with actions to reduce plastic waste are understood as we work towards the manifold economic, environmental and health objectives of the SDGs. Identifying and quantifying these effects could help to mitigate unintended consequences and provide additional incentives that could be used to advocate and propel action on recycling and reuse targets. Research suggests that circular economy strategies could provide co-benefits for reducing climate change and increasing employment (*13, 14*). Yet, the potential opportunities and risks for human health remain poorly understood; scientific evidence is scarce and health indicators are not integrated into circular economy policy monitoring(*4*).

Opportunities to improve health outcomes may plausibly be derived from reducing the impacts of a linear economy of plastics. For example, increased recycling and reuse would reduce the need for raw material extraction required by virgin plastic production and would circumvent incineration at the end of the packaging life cycle. These processes cause air pollution linked to respiratory diseases, and lead to greenhouse gas emissions which in turn increase heat related morbidity and mortality due to climate change(*15*). Conversely, these circular strategies could also pose health risks associated with occupational chemical and contaminant exposure during recycling and remanufacturing processes (*4*). Direct consumer exposure to unintentionally added chemicals in recycled plastics, or contaminants during reuse, may also be a concern, particularly in the absence of strict regulation and careful hygiene management (*4, 16*). Whilst some evidence of these health effects exists, it is often limited by challenges of attribution or restricted to discrete stages of the packaging life cycle (*15*).

To gain a more comprehensive understanding of possible health co-benefits and trade-offs of circular strategies, methods based on systems thinking are required. The European Commission endorses Life Cycle Assessments (LCAs) as the current most effective framework for assessing environmental impacts associated with products (*17*). LCA can also provide estimates of health effects, calculated in Disability-Adjusted Life Years (DALYs). This estimate is the sum of human morbidity and mortality due to climate change, ozone formation and depletion, ionising radiation, human toxicity, and water scarcity **(Figure 1)**, calculated by applying conversion factors based on the latest scientific evidence to a complete inventory of the material and energy inputs and outputs of a product and its life cycle processes (*18*). This unique assessment of the entire life cycle allows circular versus linear economy health trade-offs to be analysed across plastic packaging production, transport, consumer use and disposal.

**Fig. 1.**
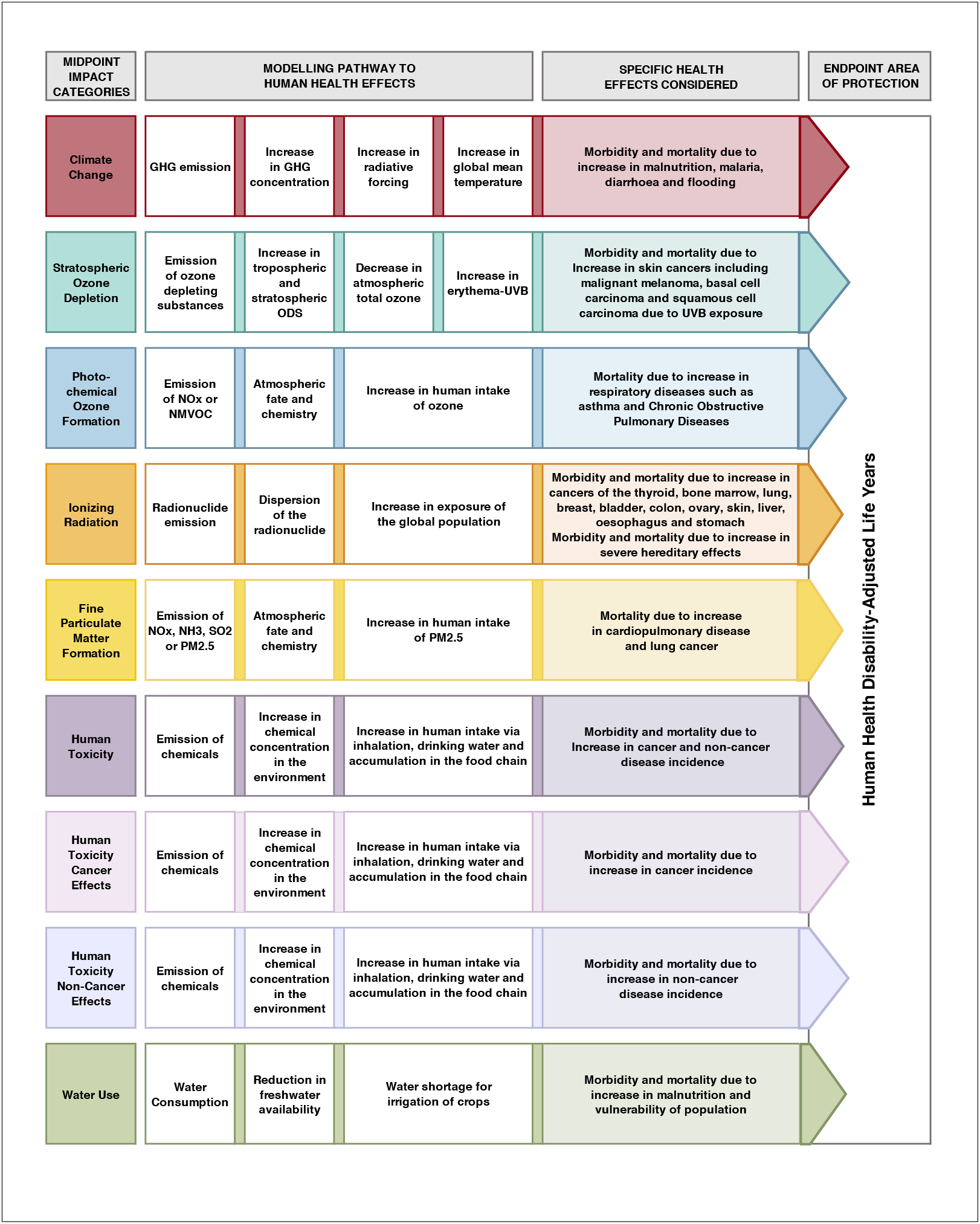
Modelling human health effects in life cycle assessments. Life cycle assessment conceptual modelling process of health impact pathways from midpoint health-related categories to endpoint Human Health Disability-Adjusted Life Years (DALYs), according to ReCiPe 2016 life cycle impact assessment method. Authors’ own: Adapted from ReCiPe 2016 v1.1 Report 1: Characterization. 2017 (*90*).

LCAs are increasingly used to examine the relative effects of circular economy strategies and have been conducted for a range of single use plastic and consumer-level packaging formats, some of which report health-related impacts (*19*–*21*). These studies have been reviewed to better understand the environmental performance of recycling and reuse options and to support policy decisions (*22*–*24*) but not yet for their contribution to understanding human health effects.

We systematically review and meta-analyse evidence from LCA studies to determine the potential value of these methods for public health research on the circular economy of plastic packaging, and to provide a quantitative analysis of possible human health risks and opportunities associated with recycling and reuse across the packaging life cycle in selected packaging types **(Figure 2)**. The two primary objectives of this paper are:

**Fig. 2.**
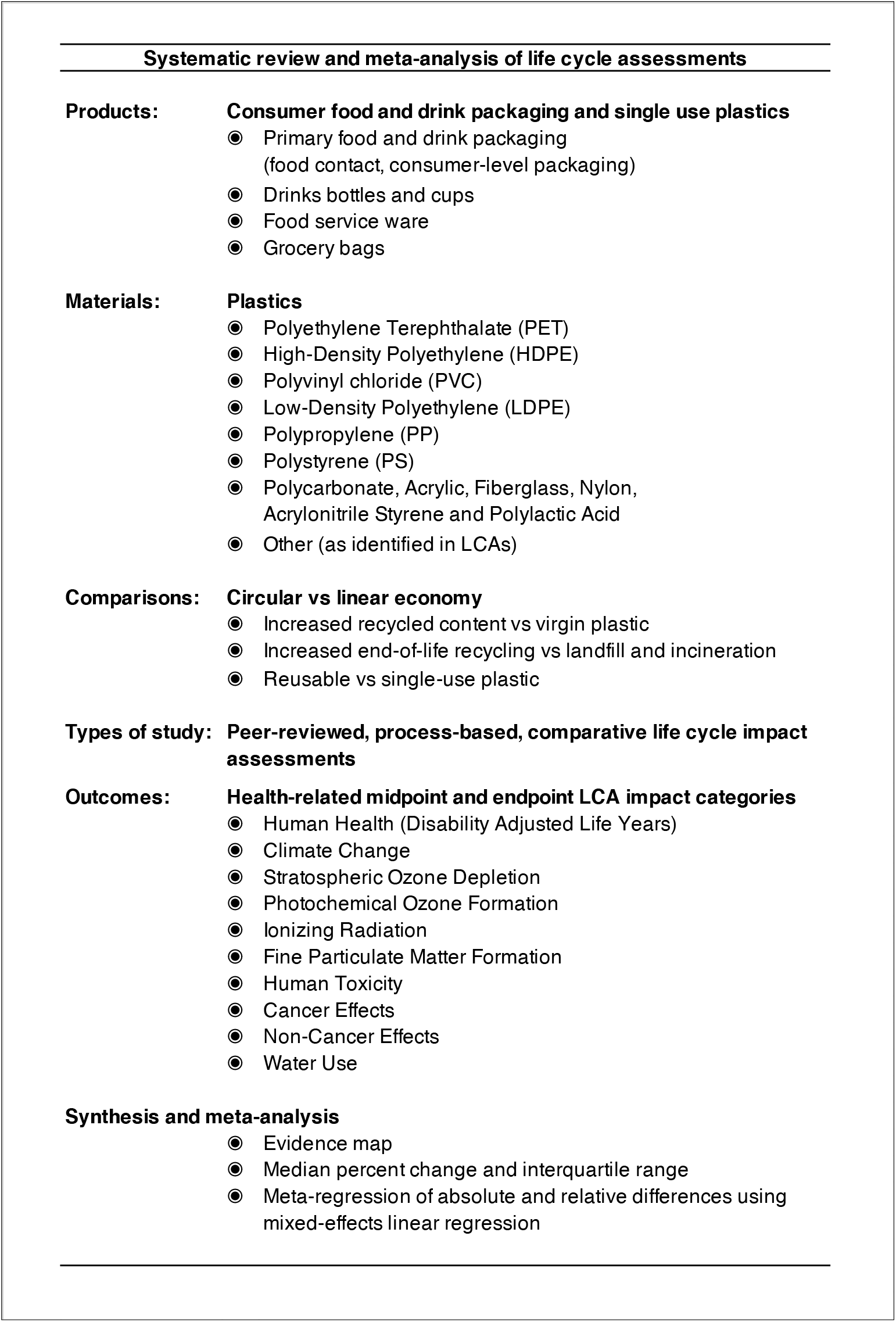
Summary of systematic review inclusion criteria and meta-analysis. Life cycle assessment (LCA) studies were systematically screened against pre-defined eligibility criteria. Studies were required to meet at least one of the inclusion criteria under each of the sub-headings. Strategy for synthesis and meta-analysis of data extracted from LCAs included an evidence map, summary statistics and regression.

Objective 1: To identify trends and gaps in the use of health-related indicators in life cycle assessments of plastic primary food and drink packaging, food service ware and grocery bags that compare virgin and single use plastic packaging to recycling and reuse options

Objective 2: To quantitatively meta-analyse the possible human health effects of circular economy scenarios according to LCA evidence, compared to a linear economy of plastic primary food and drink packaging, food service ware and grocery bags

## Results

### Characteristics of included studies

Our systematic literature search retrieved 10,499 unique records from scientific databases. After screening by title and abstract we included 532 records for full text screening, of which 30 were included in our review **(Figure 3)**. Reasons for exclusion were primarily related to study design and a lack of recycling or reuse comparison. Searching reference lists of included studies and contact with the authors resulted in eight additional studies for inclusion. The updated database search, on the 22^nd of^ July 2021, produced seven new eligible studies and grey literature searches returned four, resulting in a total of 49 LCA studies included our review.

**Fig. 3.**
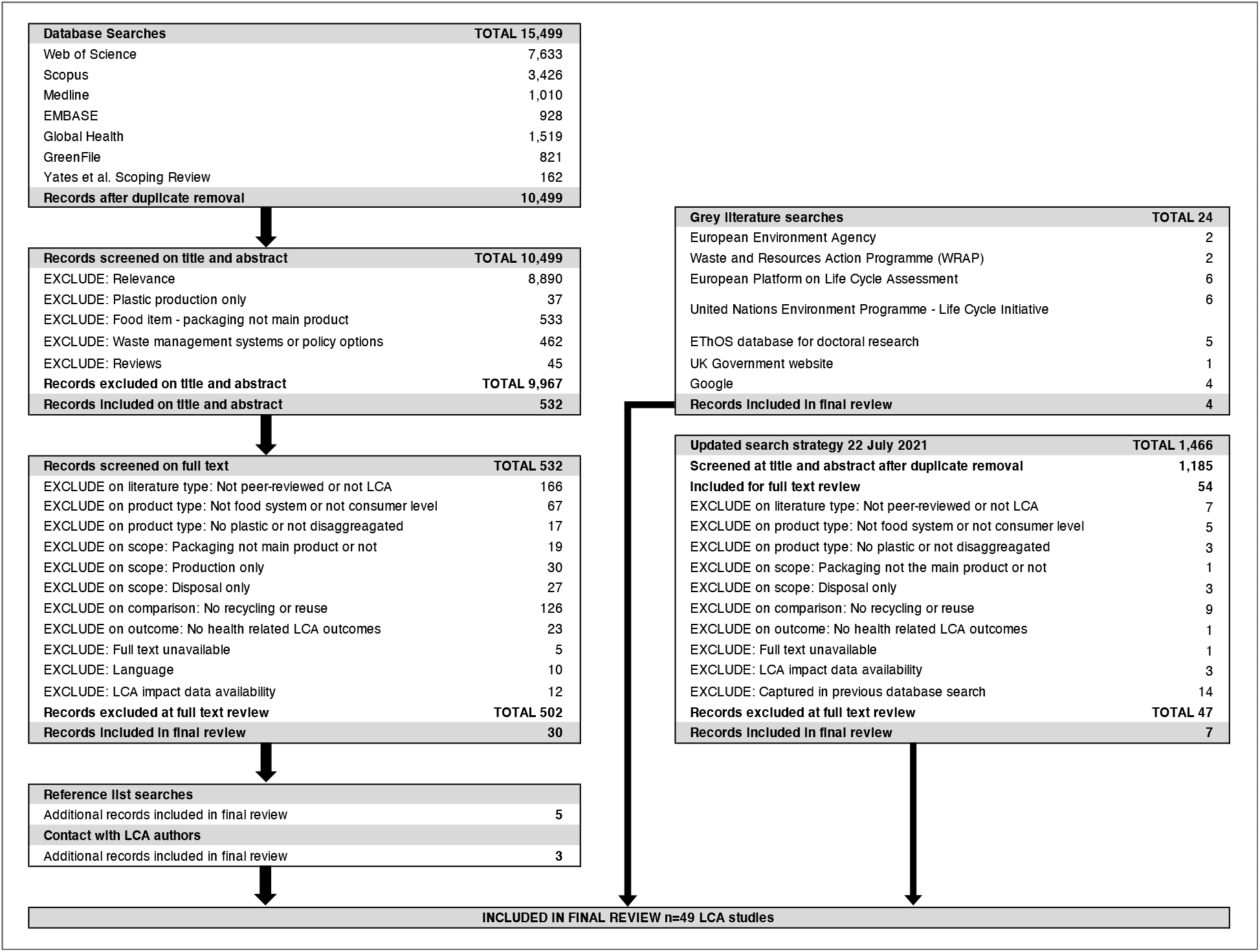
PRISMA flow chart. Preferred Reporting Items for Systematic Reviews and Meta-Analyses (PRISMA) flow chart showing the study selection process for inclusion of life cycle assessments in the systematic review according to pre-defined eligibility criteria.

All included studies were published in the last 20 years, most since 2010, with the highest number of annual publications in 2020-2021. Evidence from high-income countries in Europe and the United States of America predominated (n=44 studies), the remaining studies focused on product systems in Thailand (n=4) and China (n=1).

Within studies, comparisons focused predominantly on recycling (n=42), 15 of which included a calculation of recycled content compared to virgin material and 36 studies compared end-of-life recycling to incineration and/or landfill. Reusable products were compared to single use products in 17 LCAs. Studies most frequently assessed drinks bottles (n=15), followed by primary food packaging (n=11). Cups and service ware featured equally (n=9 studies each) and fewer studies considered grocery bags (n=7), two studies considered both bottles and packaging. These products were made of various types of plastics, the most frequent being polyethylene terephthalate (n=27), polypropylene (n=22) and polystyrene (n=16).

The impact assessments in each study was almost exclusively conducted at midpoint level; only one study considered the endpoint area of protection, Human Health in DALYs. All studies assessed Climate Change (n=49), but fewer calculated other midpoint indicators, such as Cancer and Non-Cancer Effects (n=6 and n=4 respectively). The frequencies of studies cross-referenced by circular economy strategy, product type and LCA impact assessment categories are presented as a static map **(Figure 4)**. The <u>evidence map</u> is also available online and can be interactively filtered by type of plastic, publication date and location. Complete data extraction and all meta-analysis calculations are available in **Data S1-11**.

**Fig. 4.**
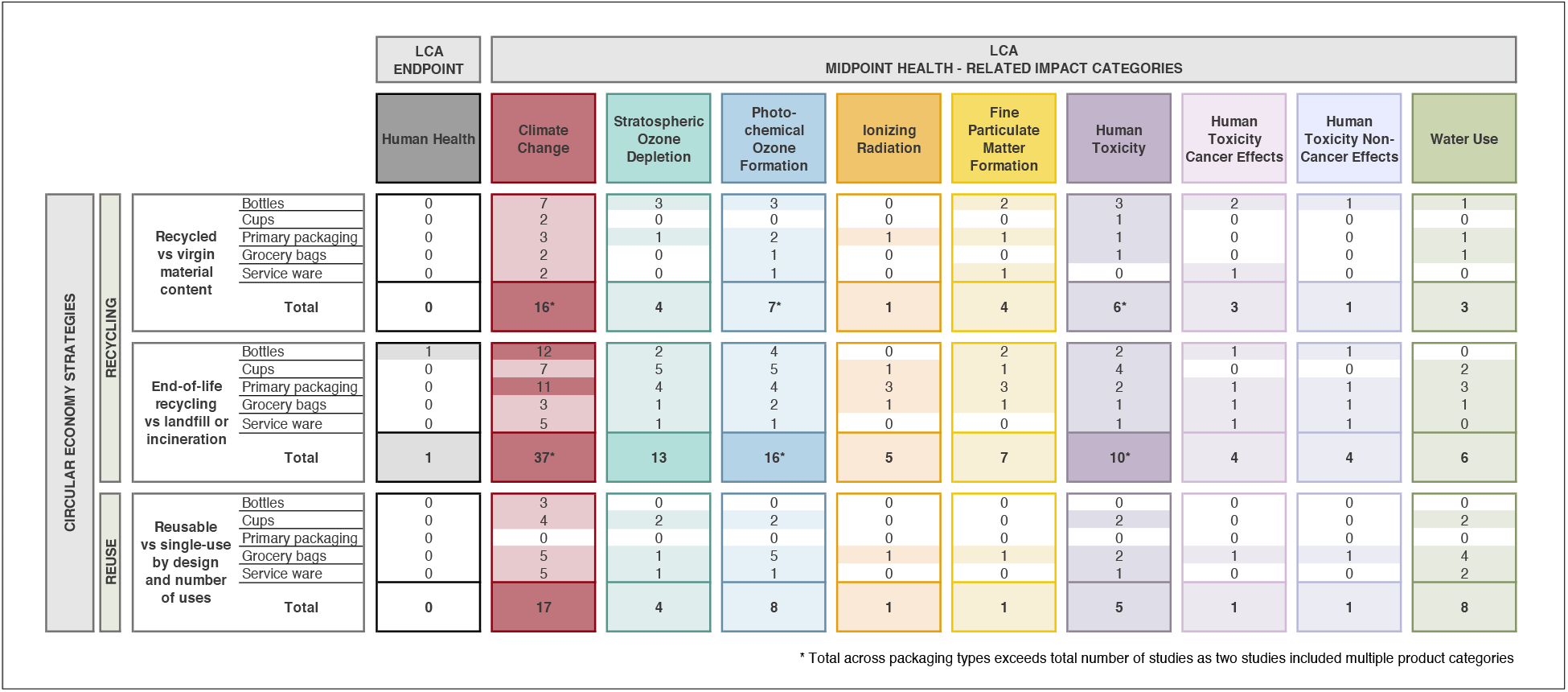
Static map of LCAs on plastic food and drink packaging recycling and reuse. Frequency of life cycle assessment (LCA) studies included in the systematic review, cross tabulated by: (A) the specific circular economy comparison categories considered in each study: recycled vs virgin material content, end-of-life recycling vs landfill or incineration, reusable vs single-use by design and number or uses; (B) the product type: bottles, cups, primary food packaging, grocery bags or service ware and (C) the health-related LCA impact categories calculated in each study.

### Reporting quality and suitability for meta-analysis

Our appraisal of included studies indicated that the majority of included LCAs modelled the packaging life cycle comprehensively **(Figure S1)**, though the consumer use phase was rarely included in recycling comparisons: most studies stated that they assumed negligible or equal impacts during this phase in the context of their comparison. Two thirds of studies provided evidence of sufficient data for all modelled life cycle stages, either providing the actual data or detailing and referencing the sources.

None of the included studies explicitly set out to determine health impacts of recycling and reuse in their stated goals though many used impact categories such as Human Toxicity and Cancer Effects that only link to Human Health as the endpoint area of protection, rather than the environmental area of protection ‘Ecosystems’. Most studies provided a detailed description of the functional units with weight measurements (88%), though fewer were explicit about the method for accounting for recycling and reuse (71%), and 8% did not provide the specific impact assessment method used.

47 studies reported total life cycle impacts, the other two presented no overall impact data but instead provided results for each stage of the life cycle with multiple scenarios. Half the included studies presented disaggregated results for at least some of the life cycle stages modelled that were relevant to our review question.

### Health effects of increasing recycled content of plastic packaging

We identified 15 LCAs that included recycled plastic content comparisons in food packaging (*20, 25*–*38*). In fixed scenarios of packaging made of 100% recycled plastic compared to 0% recycled plastic **(Figure 5)**, ten comparisons from seven studies showed consistent reductions in health-related midpoint impacts including Climate Change, Photochemical Ozone Formation, Particulate Matter Formation, and Cancer Effects for 100% recycled plastic. Within each of these categories, the range of values was wide, with the median indicating average impacts a quarter to half the magnitude of those of virgin packaging. The median reduction for Ozone Depletion was the smallest at 7.22% (comparison points (cps)=4; IQR= -21.78% to -7.11%), and Human Toxicity was calculated only once, showing a 57.14% reduction in impacts relative to virgin packaging. Water Use was the only assessed impact where the direction of the relationship was unclear: the median showed a 7.03% increase in impact for 100% recycled plastic but one estimate indicated a 20% reduction (cps=3; Median=7.03%; IQR= -21.52% to 9.50%). There was a very small increase in Non-Cancer Effects from 100% recycled plastic, but this estimate came from a single study.

**Fig. 5.**
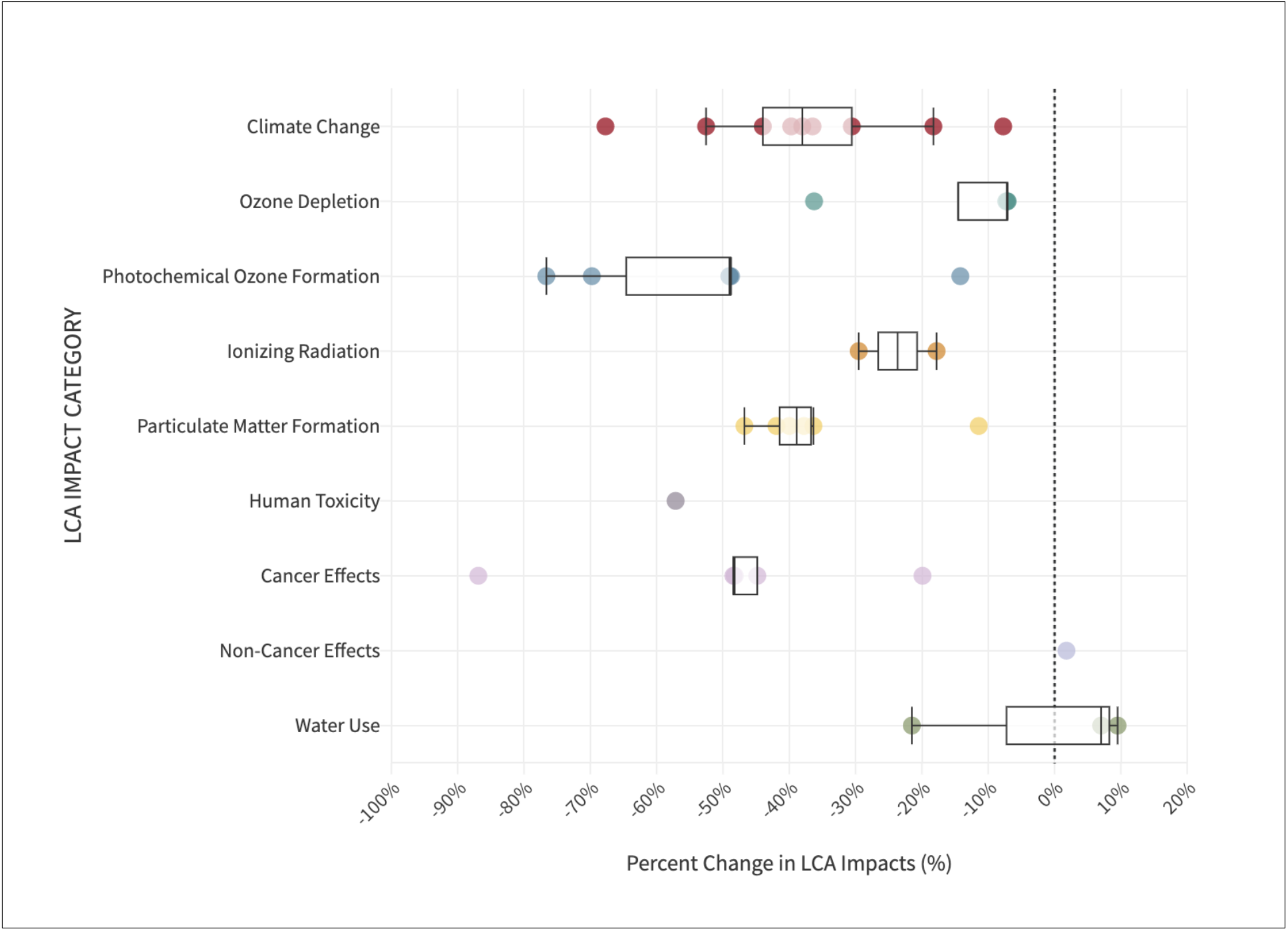
Relative health-related impacts of 100% recycled plastic content. Percent change in health-related impacts, comparing 100% recycled plastic content products to virgin plastic products. Values calculated for each comparison found within LCA studies are represented with dots and summarised using box plots of the median and interquartile range across data points for Climate Change, Ozone Depletion, Photochemical Ozone Formation, Ionizing Radiation, Particulate Matter Formation, Human Toxicity, Cancer Effects, Non-Cancer Effects and Water Use.

Meta-regression analysing the effects of increasing recycled content, relative to baseline 0% recycled plastic, showed a linear relationship with decreasing Climate Change impacts and Total DALYs based on 41 packaging comparison points extracted from 13 studies (**Figure 6**). 21 comparison points were based on bottles, six on service ware, five on cups, five on grocery bags and four on food packaging. On average, relative to virgin plastic packaging, there was an additional 161kg decrease in Climate Change impacts (kg CO2 equivalents) for every 10% increase in recycled content in 1 metric tonne of food and drink plastic packaging (Coefficient=-16.0978; 95% CI= -20.64423 to -11.55136; p<0.0001). This equates to a 0.000149 decrease in DALYs as a result of climate change.

**Fig. 6.**
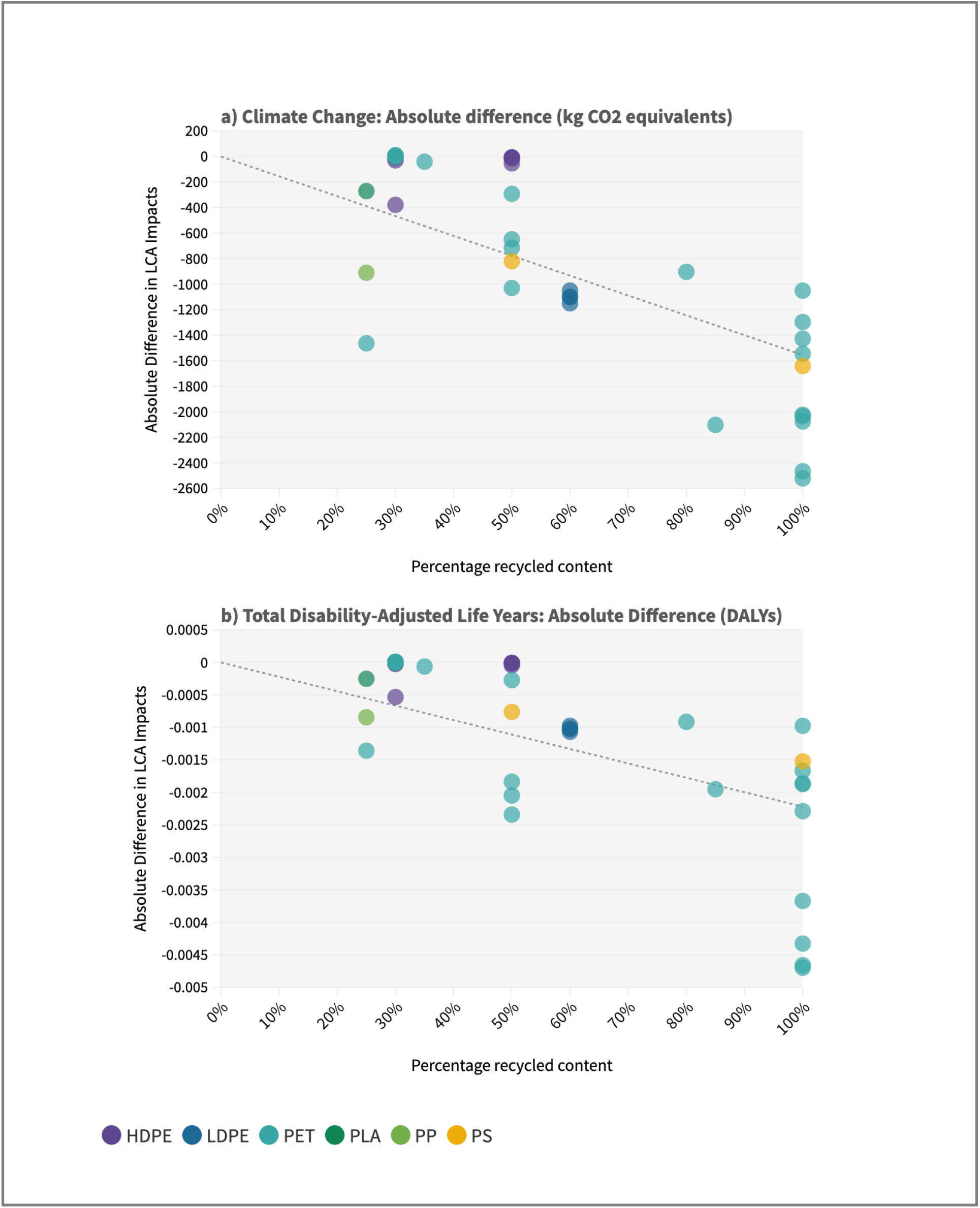
Meta-regression of the effects of increasing recycled plastic content on Climate Change and Total DALYs. Scatter plot for mixed-effects linear regression with percentage recycled content as the predictor variable and the response variables: **(A)** absolute difference in Climate Change impacts (kg CO_2_ equivalents) relative to virgin plastic packaging and **(B)** absolute difference in Total Disability-Adjusted Life Years (DALYs) based on multiple LCA health pathways, relative to virgin plastic. The key shows the colour coding of comparison points by plastic material type: High-density Polyethylene (HDPE), Low-density Polyethylene (LDPE), Polyethylene Terephthalate (PET), Polylactic Acid (PLA), Polypropylene (PP), Polystyrene (PS).

There was insufficient data on other midpoint impact categories to create separate regression models. However, for each comparison point, all provided midpoint indicators were converted to DALYs and summed to give a Total DALYs estimate for each comparison. Estimates of Total DALYs were therefore based on different numbers of midpoint impacts. All included the Climate Change pathway, and some included other midpoint impacts such as Ozone Depletion, Photochemical Ozone Formation, Ionizing Radiation, Particulate Matter Formation, and Human Toxicity impacts depending on the study **(Data S3)**. The trend for Total DALYs remained in favour of increasing recycled content. On average, relative to virgin plastic packaging, there was an additional 0.000196 decrease in Total DALYs for every 10% increase in recycled content in 1 metric tonne of food and drink plastic packaging (Coefficient -0.0000196; 95% CI= -0.0000269 to -0.0000124; p<0.0001). This equates to nearly a day of healthy life gained for every tonne of packaging made from 100% recycled plastic as opposed to virgin plastic (0.72 days).

Overall, reductions in Total DALYs were largely due to reductions in Climate Change impacts, though in some cases Climate Change effects contributed to increasing DALYs **(Figure S2)**. Reductions in Particulate Matter Formation contributed to reductions in Total DALYs more than the Climate Change effects in six packaging comparison points and Human Toxicity appeared generally to support reductions in DALYs though increased them in three comparison points.

Two studies could not be included in quantitative meta-analysis because the recycled content comparison was part of a mixed strategy scenario, but both showed decreased impacts relative to other scenarios (*25, 28*).

### Health effects of increasing end-of-life recycling of plastic packaging

End-of-life recycling was compared to incineration and/or landfill in 36 studies (*19*–*21, 25*–*29, 31, 33, 38*–*63*). The only study to report on Human Health DALYs calculated a reduction of 0.9 *μ*DALYs (−28%) for 100% recycling of 4 PET wine bottles compared to a scenario of equal landfill and incineration (*19*).

In discrete scenarios, studies that modelled 100% recycling compared to a reference 100% incineration (n=19 studies, 92 comparison points) or 100% landfill (n=12 studies, 52 comparison points) showed mixed results for health-related midpoint impacts with wide ranging values across studies **(Figure 7)**. All studies that compared 100% recycling to 100% landfill reported percent decreases in Climate Change, Ozone Depletion, Ionizing Radiation, Particulate Matter Formation, Cancer Effects and Water Use, though only Climate Change was based on more than four comparison points (cps=52; Median= -40.53%; IQR= -54.83% to -22.09%). Effects were mixed for Photochemical Ozone Formation (cps=22; Median= -15.68%; IQR= -53.98% to -5.80%), Human Toxicity (cps=19; Median = -24.57%; IQR=-45.59% to -12.07%) and Non-Cancer Effects (cps=4; Median= -58.50%; IQR= -71.00% to -28.00%) with some studies showing relative increases in impacts, though the median presented an average reduction in each impact category. Results were more variable in studies that compared 100% recycling to 100% incineration. Whilst studies consistently showed reductions in Climate Change (cps=79; Median= -56.59%; IQR= -68.78% to -36.89%) and Non-Cancer Effects (cps=3; Median= -79.45%; IQR= -84.16% to -79.22%), increases in impacts were seen for Ionizing Radiation (cps=10; Median=98.34%; IQR=31.91% to 183.33%) and all other indicators showed both relative decreases and increases in impacts compared to incineration, resulting in median percent increases in Ozone Depletion (cps=17; Median=27.27%; IQR= 7.18% to 378.57%), Particulate Matter Formation (cps=11; Median=85.71%; IQR= -29.38% to 90.91%) and Water Use (cps=17; Median=16.00%; IQR= -12.77% to 81.13%). Human Toxicity was the only indicator for which the median and mean were not aligned on the direction of effect: the median showed decreased impacts and the mean an increase.

**Fig. 7.**
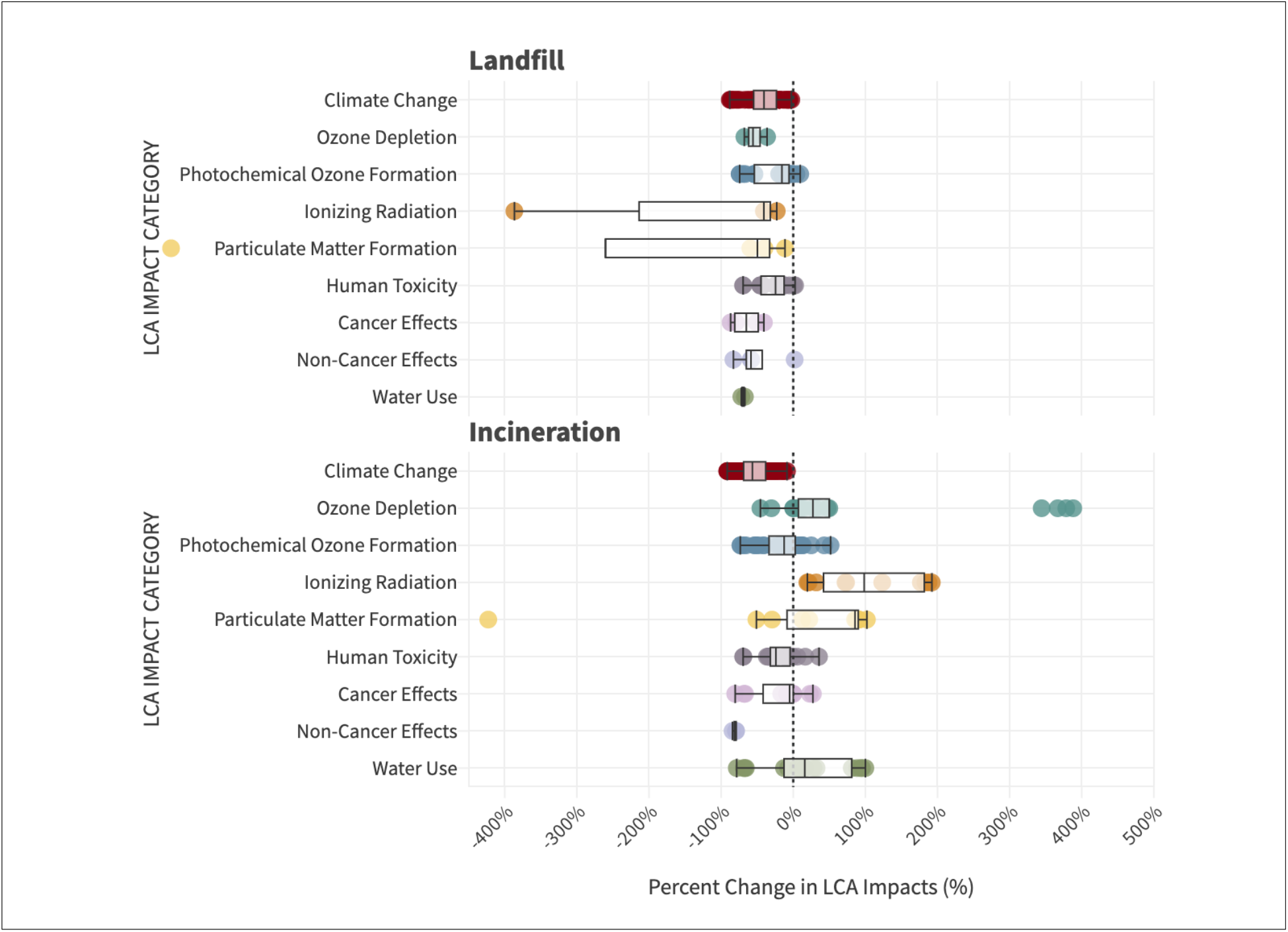
Relative health-related impacts of 100% end-of-life plastic recycling. Percent change in health-related impacts, comparing 100% recycling to **(A)** 100% landfill and **(B)** 100% incineration. Values calculated for each comparison found within LCA studies are represented with dots and summarised using box plots of the median and interquartile range across data points for Climate Change, Ozone Depletion, Photochemical Ozone Formation, Ionizing Radiation, Particulate Matter Formation, Human Toxicity, Cancer Effects, Non-Cancer Effects and Water Use.

We used mixed-effects linear regression to meta-analyse the effects of increasing end-of-life recycling compared to a baseline 100% landfill and/or incineration. We included 179 comparison points from 27 studies; 65 comparison points were based on food packaging, 61 on bottles, 23 on service ware, 15 on cups and 15 on grocery bags. Relative to landfill and/or incineration, there was an average additional 227kg decrease in Climate Change impacts (kg CO2 equivalents) for every 10% increase in end-of-life recycling rate of 1 metric tonne of food and drink plastic packaging (Coefficient=-22.69839 95% CI= - 27.22419 to -18.17259; p<0.0001) **(Figure 8)**. This equates to 0.000210656 DALYs mediated only by climate change.

**Fig. 8.**
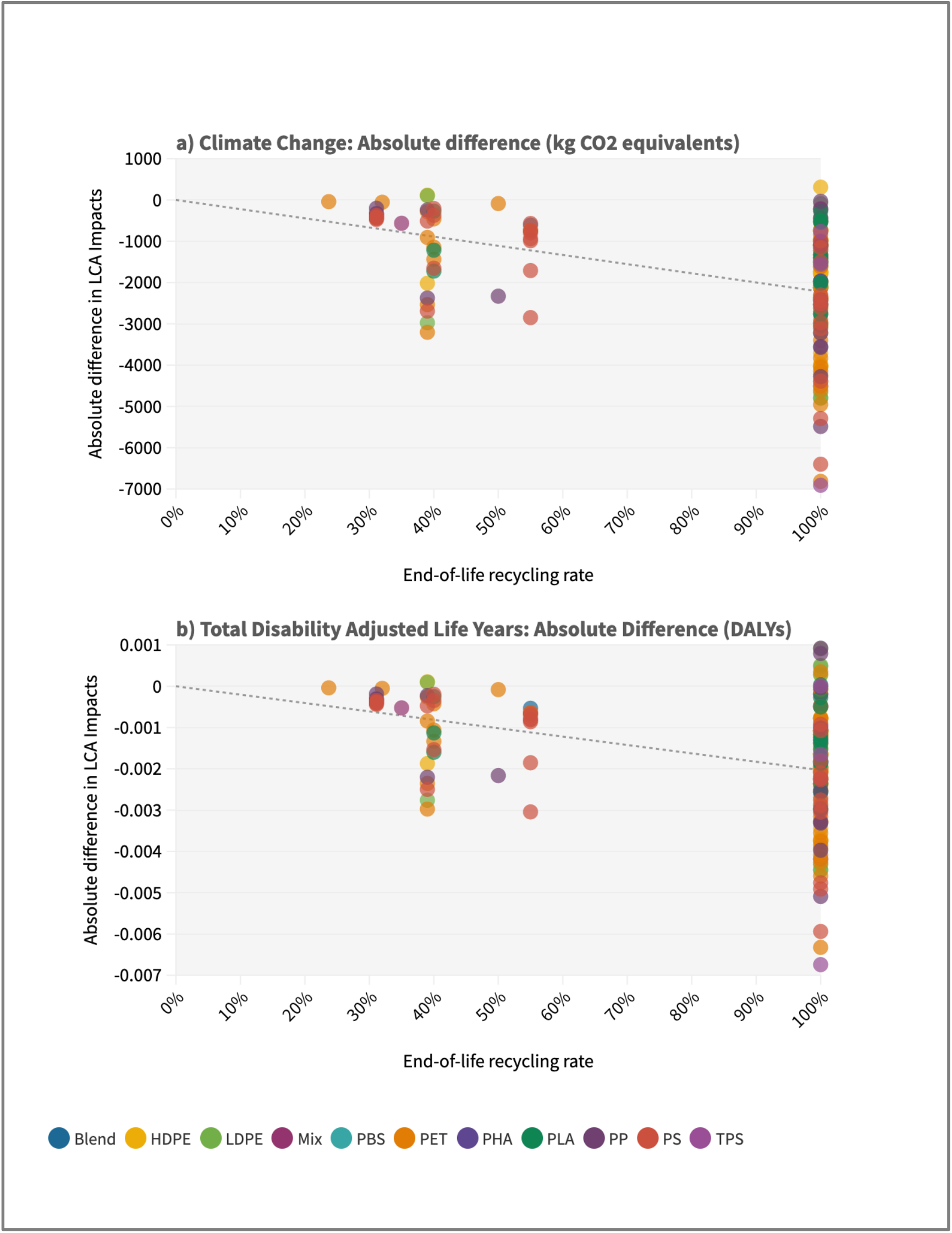
Meta-regression of the effects of increasing end-of-life recycling on Climate Change and Total DALYs. Scatter plot for mixed-effects linear regression with end-of-life recycling rate as the predictor variable and the response variables: **(A)** absolute difference in Climate Change impacts (kg CO_2_ equivalents) relative to 0% recycling and **(B)** absolute difference in Total Disability-Adjusted Life Years (DALYs) based on multiple LCA health pathways, relative to 0% recycling. The key shows the colour coding of comparison points by plastic material type: Compound plastics (Blend), High-density Polyethylene (HDPE), Low-density Polyethylene (LDPE), Multiple components made of different plastics (Mix), Polybutylene Succinate (PBS), Polyethylene Terephthalate (PET), Polyhydroxyalkanoates (PHA), Polylactic Acid (PLA), Polypropylene (PP), Polystyrene (PS), Thermoplastic starch (TPS).

Total DALYs based on all midpoint indicator estimates provided for each LCA comparison point remained in favour of increasing recycling. On average, relative to 0% recycling there was an additional 0.000211 decrease in Total DALYs for every 10% increase in recycling of 1 metric tonne of food and drink plastic packaging (Coefficient - 0.0000211; 95% CI= -0.000026 to -0.0000163; p<0.0001). This equates to nearly a day of healthy life gained for every tonne of packaging recycled instead of incinerated and landfilled (0.77 days). Climate Change impacts consistently contributed to decreasing DALYs though contributions from Human Toxicity measures, Particulate Matter Formation and Water Use were mixed, and in some cases increased DALYs in recycling scenarios **(Figure S3)**.

Eight LCAs could not be included in meta-regression because of challenges with comparability of data and scenarios. The specific reasons are listed in **Data S4**. However, these studies also predominantly reflected that increasing recycling could reduce health-related impacts relative to landfill and incineration.

Differences in impacts for recycled content and end-of-life recycling comparison points were generated almost entirely from emitting processes and credits awarded for avoided virgin material, at the production and waste management stages of the packaging life cycle. Where studies provided disaggregated data, some calculated differences in transport stages, as a result of different collection systems, but this only accounted for 1-12% of the total difference in impacts.

### Health effects of reusable plastic packaging

Reusable products were compared to single use products in 17 LCAs (*21, 25, 28, 30, 35, 41, 43, 56*–*58, 64*–*70*). Studies comparing packaging designed to be reused with packaging designed for single use, based on both products being used just once (n=6 studies; 32 comparison points) demonstrated almost universal and often large relative increases in all health-related impacts for reusable packaging **(Figure S4)**. Relative to single use, reusable packaging showed a median increase of 852.17% (cps=32; IQR=382.60% to 1260.93%) in Climate Change impacts, increased Ozone Depletion by 684.87% (cps=20; IQR= 237.33% to 2610.98%), Photochemical Ozone Formation by 969.07% (cps=30; IQR= 207.69% to 1690.54%), Human Toxicity by 447.59% (cps=12; IQR= 219.84% to 1196.52%) and Water Use by 4689.27% (cps=4, IQR=673.21% to 8780.39%).

The reuse mixed-effects regression model used percent change in impacts data from 13 studies with 97 packaging comparison points; 45 comparison points were based on grocery bags, 32 on cups, 13 on bottles and 7 on service ware. Compared to the impact of single-use packaging used once, the percent change in Climate Change emissions of reusable packaging decreased on average by 15.05 percentage points for every additional use (Coefficient=-0.1504829; 95% CI= -0.3118651 to -0.0108993; p=0.068). In brief, whilst the impact of reusable packaging is initially relatively higher than single use packaging, the relative difference becomes smaller with each additional use until equivalent and eventually smaller impacts are generated. The break-even point for Climate Change impacts, (where percent change=0) occurs at 29.94 uses, meaning that on average reusable packaging must be used at least this many times to equal or reduce Climate Change impacts and climate associated DALYs, compared to single use packaging.

Due to the limited number of studies, formal sub-group analysis by product and material type was not possible, though no visible patterns were identified in the scatter plot **(Figure 9)**. Other impact categories did not meet the threshold of ten studies needed for formal regression analysis but scatter plots with a line of best fit are shown in **Figure S5** and show linear relationships between increased reuse and reduced Ozone Depletion, Ozone Formation, Human Toxicity and Water Use impacts relative to single use packaging.

**Fig. 9.**
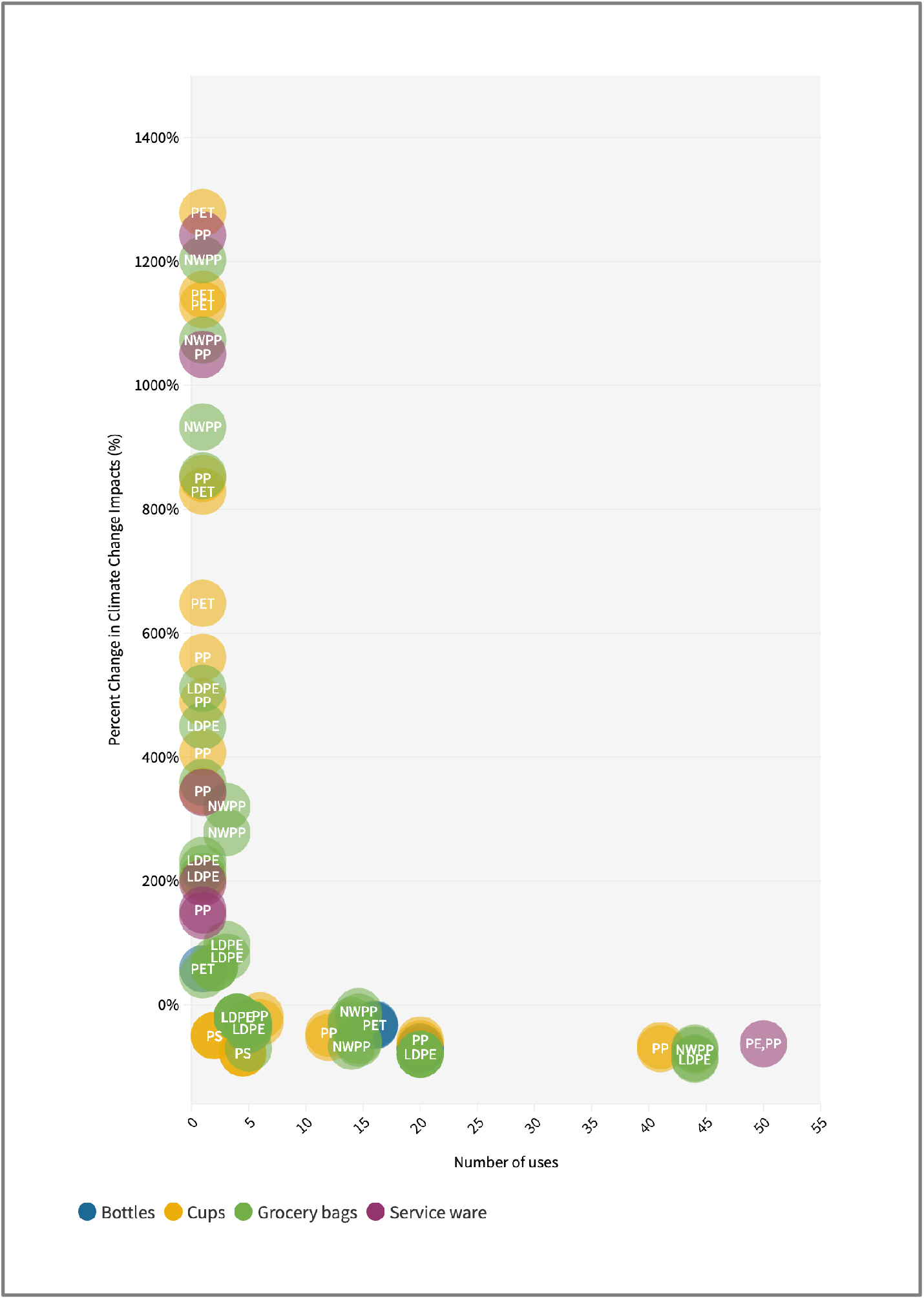
Meta-regression of increasing reuse compared to single use. Scatter plot for mixed-effects linear regression with number of uses of the reusable plastic packaging as the predictor variable and the percent change in Climate Change impacts relative to the single use counterpart packaging. The specific plastic material type of the reusable product is written over corresponding data points, a full list is provided in **Data S11**. Data points are colour coded on product type: Blue=bottles, yellow=cups, green=grocery bags, purple=service ware. A full list of material and product types are available in Data S11. For ease of viewing the y axis has been capped at 1500% but there are six data points above this point, all are based on one use, 5 of which refer to PLA cups had between 1631% and 7450% higher Climate Change impacts and one PET cup which had 2238% higher Climate Change impacts than the respective single use comparison. Abbreviations: Low-density Polyethylene (LDPE), Non-woven Polypropylene (NWPP), Polyethylene (PE), Polyethylene Terephthalate (PET), Polypropylene (PP).

Five studies in this review were not included in the quantitative reuse meta-analysis; two had no single-use baseline and three had no absolute impact data but provided their own break-even points. For Climate Change impacts, these were 4 times for reusable plastic cutlery (*67*), 2-4 times for takeaway food containers (*68*), <3-7 times for drinks cups (*70*).

## Discussion

Circular economy strategies including recycling and reuse are primarily designed to reduce waste and pollution. We found evidence for possible co-benefits to human health as a result of these strategies in plastic food and drink packaging. This was supported by strong evidence for a linear trend between both increasing percentage recycled content and increasing end-of life recycling rate with decreasing DALYs relative to virgin plastic and incineration or landfill waste management. Savings in DALYs were mainly a result of reduced greenhouse gas emissions, which contribute to climate change and associated morbidity and mortality, though other pathways also contributed. Scaling up our results, for every 10% of the current annual global food and drink packaging waste (*2, 3*) recycled instead of landfilled or incinerated, we could save around 10,400 DALYs, though we identified critical gaps in this evidence. Despite overall improvements for human health, some studies reported increased risks from Human Toxicity, Particulate Matter Formation and Water Use, suggesting possible trade-offs that are currently under-explored. We found that reusable plastic packaging can reduce health impacts associated with single use plastics, but on average across packaging formats, these reductions only occur with frequent reuse, not reflected in current consumer behaviour. Life cycle assessments that compare circular versus linear economies of plastic primary food and drink packaging represent an emerging body of literature. We identified opportunities for future research to build on LCA methods towards a more robust and comprehensive evidence base for assessing circular economy health risks and opportunities.

### Meta-analysis results in the research context

Our estimates of possible human health co-benefits of recycling plastic packaging were smaller than those calculated for some other environmental strategies, for example the health co-benefits estimated for more sustainable dietary choices (*71*). However, the projected growth in plastic consumption means that health effects are important to understand (*72*). An LCA of personal protective gear, made predominantly of single use plastics, was conducted for the first six months of the COVID-19 pandemic in the UK (*73*). Authors compared maximum recycling to baseline incineration and reported an absolute reduction of 68 DALYs (*73*), which on a per tonne basis was aligned with our findings for plastic food and drink packaging.

Meta-analysis results for midpoint health-related indicators broadly aligned with existing studies evaluating the environmental impacts of packaging recycling and reuse. A review by *Vendries et al*. of packaging and food service ware compared higher versus lower levels of recycled content in packaging made of any plastic or other material (*23*). Our findings were similar in that most comparisons showed reduced impacts with increased recycled content. Some increased impacts were seen in three newer studies included in our review but other studies from the same time-point did not. Reasons for these differences are unclear and highlight possible trade-offs that remain critically underexplored.

Our findings on the absolute reduction in Climate Change impacts as a result of end-of-life recycling are largely in line with a broader study by *Lau et al*. on all plastic packaging using stochastic modelling (*72*). This study reported a reduction of 1.9tCO_2_ equivalents per metric tonne of packaging recycled, relative to virgin plastic production (*72*). Using the ReCiPe 2016 conversion factor for Climate Change impacts, this would equate to a reduction of 0.0017632 DALYs per tonne of packaging. Our meta-analysis produced slightly higher estimates, showing average reductions of 2.3tCO_2_ equivalents per tonne and a 0.00208 decrease in Total DALYs relative to incineration or landfill. Differences are likely because our meta-analysis is based on the distribution of material types found in the literature, whereas this modelling study was based on market share. In addition, our estimate of DALYs incorporated multiple health pathways using all available indicator data from included studies.

For reusable packaging, our meta-analysis suggests a break-even point of 30 uses, higher than reported in some other studies. Two other reviews of life cycle assessments comparing reusable products to single use have been conducted (*24, 74*). One was not restricted to plastic materials and included a broader range of packaging types, including transport packaging, reporting break-even points between 2-20 uses(*74*). The other was restricted to single use plastic carrier bags and their alternatives, the break-even point for plastic reusable bags was between 5-20 uses (*24*). No formal quantitative meta-analysis was conducted in either review, so figures represent a range of break-even points reported in individual studies. Our results are higher than these figures, probably because we specified single use plastic as the baseline for all our comparisons. Whilst highly pollutive, single use plastic can perform relatively well in other environmental categories so the break-even point for reusables will be higher. The second review did restrict comparisons to those with a single use plastic baseline but the difference in weight and production impacts between reusable and single use bags is likely to be less than for some other packaging types. Our analysis reflects an average across packaging types and plastic materials, this is likely to make the average reuse required higher because of the inclusion of comparisons such as a single use polystyrene food saver with a reusable Tupperware container, which have very different weights. We did not find enough studies to conduct a formal subgroup analysis within the meta-regression, but our conclusions are in accordance with these reviews in that the impacts of reusable products will reduce each time they are used and the break-even point is relatively high. This means that products need to be sufficiently durable, and that consumer awareness and behaviour reflects the required level of reuse (*24, 74*).

### Meta-analysis results in practice

Life cycle assessments often model impacts based on hypothetical scenarios rather than real-life recycling and reuse systems. This is useful in indicating trends for increasing circular economy activities but does not always align with technological and economic possibilities or with current consumer behaviour.

Many of the studies we included modelled impacts of using 100% recycled plastic content and 100% end-of-life recycling. However, each reprocessing degrades material quality meaning that infinite recycling is not possible. Stochastic modelling suggests that mechanical recycling could be scaled up by 2040 to process 33% of global total plastic municipal solid waste but the remainder would not be possible due to capacity for waste collection, the economics of recycling and material losses(*72*). These restrictions on expanding recycling capacity will be acute in low- and middle-income countries, particularly those where large distances and dispersed rural populations pose challenges for infrastructure and where recycling may not be a political or economic priority. This means that the reductions in health impacts that can be calculated through scenario modelling may not be realisable due to real-world external constraints on the recycling system. Further to this, recycling is modelled as if it took place in an industrial, high-income setting, which does not account for the potentially higher health risks involved in the informal recycling sector commonplace in LMICs (*75*) and the heavy reliance on international waste trade that directs waste from HICs to LMICs (*76*). The UK for example exports most of its recycling, often to countries that do not have the capacity to process it, with increasing reports of resulting environmental and health damages (*77, 78*).

Nationally representative surveys in nine European countries, including the UK, indicated that consumer reuse may be significantly lower than that required to break-even or reduce impacts compared to single use packaging (*79*). Whilst a quarter of respondents reused heavyweight plastic carrier bags more than 10 times, only 8% reused lightweight plastic bags as much. 27% and 40% of respondents reused these bags just once or not at all (*79*). Well-intentioned redesign for reuse and national policies may be having the unintended effect of increasing environmental and health risks if consumer behaviour does not correspond with the necessary level of reuse.

Both circular economy strategies have been affected by the COVID-19 pandemic. Due to hygiene and safety concerns, reusable takeaway services have not been available and recycling capacity has been restricted by lockdowns and worker illness (*80*). We cannot ignore that these circular strategies are subject to external shocks that can limit implementation and introduce new risks and opportunities for human health.

### Evidence gaps in life cycle assessments

The most important gaps in this body of literature are the small number of LCAs on this topic and the narrow geographical coverage, the inconsistent use of available health-related indicators and the current limitations of LCA methods for health research.

Given the surge of policy action to increase recycling and reuse, promoting the circular economy over the linear, we found surprisingly few LCAs that assessed these actions in primary food and drink packaging. This was particularly the case for recycled content and reusable packaging; many of these assessments were conducted several years ago and may be relying on data that has subsequently changed. The increased number of publications during the last two years suggests a growing focus on the plastics circular economy in LCAs. Endorsement by the European Commission and the rapid evolution of policy may help to fuel a larger evidence base over the coming years. Investments should be made to ensure this evidence is more inclusive of low- and middle-income countries, which were almost entirely absent in the literature we identified and yet these are the countries that will experience the greatest future growth in plastic consumption (*81*). This underscores the existing call for the European Union and China to engage in greater dialogue with low- and middle-income countries and to foster international sharing and learning on the circular economy(*12*).

All included studies considered Climate Change impacts; based on the Intergovernmental Panel on Climate Change (IPCC) Global Warming Potentials, this is considered a relatively robust impact category to model (*82*). Understanding this impact is critical in the context of the climate crisis but restricting modelling to a single impact category offers a one-dimensional view of the effects of circular economy actions, possibly belying trade-offs in other impacts such as Particulate Matter Formation and Human Toxicity, for which we found some evidence of increased risk (*83*). The lack of estimates of DALYs may be due to the high level of uncertainty in endpoint calculations, resulting from the complexity of impact mechanisms, the challenge of predicting population adaptation and dynamics, and regional differences in impacts (*82*). However, this represents an overlooked opportunity to consider the broader picture of links between circular economies and health.

LCA is a holistic tool for assessing the health effects of circular economies. However, emerging health concerns associated with specific life cycle stages are not yet captured by LCA studies. For example, epigenetic health risks from direct consumer contact with chemicals leaching from recycled plastics has been suggested (*4*) but the use phase in LCAs of recycling was excluded from modelling. At the end of the life cycle, industrialised recycling was compared to industrialised incineration and landfill, which reflects the European and North American bias in this body of literature. This does not account for the health effects of open burning of plastics that is commonplace in many other countries, potentially reaching 144 million metric tonnes of plastic burned annually by 2040 (*72*). Neither does this include the contamination risks of residing near open rubbish dumps, occupational hazards of the informal recycling sector including waste pickers, and the health effects of environmental plastic pollution. One report estimated that diseases caused by plastic and other mismanaged waste cause 400,000 to 1 million deaths every year in low- and middle-income countries(*84*) and so the health benefits of recycling may be vastly underestimated.

### Strengths and Limitations

Our meta-analysis was designed to provide an illustrative overview of potential pathways to human health risks and opportunities as a result of circular economy strategies as identified in life cycle assessments, and an indication of the possible magnitude of effect on DALYs under certain hypothetical scenarios. Results should not be interpreted as predictive of health effects but used to support further investigation of these pathways.

Standardising impacts from studies based on different LCA methods, using ReCiPe conversion factors, can result in estimates based on different assumptions. However, this method is internationally recognised and where conversion from other methods was not possible, we excluded these results from the meta-analysis to minimise the effect. ReCiPe 2008 factors likely result in lower estimates of DALYs than the 2016 updated conversion factors. The older factors were only applied where necessary when Human Toxicity impacts were not disaggregated by Cancer and Non-Cancer Effects. New LCAs in this context using the updated 2016 midpoint impacts are required. We chose to sum all health-related midpoint impacts within individual comparison points to provide a Total DALY estimate. Different numbers of midpoint categories in this calculation, according to data available, may explain some of the variability in total estimates.

The search strategy and literature selection process followed standardised procedures for conducting systematic reviews. Conducting the literature search in English carries some bias in that LCAs published only in other languages may not have been captured, though we were able to review studies in French and Spanish where identified. We did not target the search to capture industry LCA reports though it is likely that most of these would focus on carbon emissions and not add more to our analysis in identifying health effects.

Included studies were peer-reviewed or been subject to critical review and were therefore considered to be more robust, however not all reported explicitly on key elements of their methodology. In addition, 12 studies had to be excluded because of a lack of data on the impacts of our comparisons. In some cases, this was because studies were old and the data were no longer available. This highlights the need for transparent and comprehensive data reporting in published LCAs.

We found insufficient packaging comparisons to conduct formal subgroup analysis of key attributes that would likely affect the results. Of particular interest might be sub-group analyses by packaging format and plastic material types. Accounting for differences in methodological choices in individual studies would also provide more robust estimates as identified in a critical review of LCAs of cups(*85*). For example, analyses based on the inclusion of specific life cycle stages in the system boundary, the inclusion of secondary packaging in the inventory, whether credits are given for energy recovery from incineration and landfill comparisons, the method of allocating benefits from recycling and the specific impact assessment method used. As the field grows, it would be useful to determine the influence of these factors on meta-analysis results.

### Recommendations

Circular economy strategies, including recycling and reuse, can offer co-benefits for human health through small reductions to the DALYs associated with a linear economy of plastic food and drink packaging. There is also some evidence of increased health risks though these remain critically underexplored in the literature. LCAs are a unique tool that can be used to examine multiple health indicators using a systems approach. However, many potential health pathways are not accounted for in LCAs, so results may underestimate the human health opportunities in circular economies. Building on LCA methodological capacity to support interdisciplinary research on health outcomes would facilitate a more comprehensive understanding of possible co-benefits and trade-offs associated with sustainable initiatives.

To develop the LCA evidence base for supporting understanding of health co-benefits and trade-offs associated with circular economy strategies to reduce food packaging waste:

- Conduct LCAs of food and drink packaging with circular economy comparisons, based on current technologies, systems, data and conversion factors
- Conduct international studies and foster capacity development for conducting LCAs in LMICs that will yield more representative evidence facilitating understanding of global systems and impacts
- Encourage greater methodological reporting transparency and provision of data inputs
- Encourage greater use of health-related impact categories, particularly Particulate Matter Formation, Human Toxicity, and Human Health DALYs in LCAs
- Develop LCA data pools and methodological capacity for modelling health impacts from consumer use and end-of-life open burning, landfill, and pollution
- Consider hybrid approaches, using LCA methods combined with other forms of modelling, allowing for the inclusion of more diverse evidence such as qualitative data

To aid future meta-analyses of LCA data:

- Develop guidance for conducting meta-analyses of LCAs, including tools and advice for harmonising differences in LCA methodological choices
- Develop a quality appraisal tool that can be used by non-specialists to evaluate evidence from LCAs to support accurate interpretation of results

Sensitising policy to health risks and opportunities:

- Interdisciplinary design of health-sensitive circular economy policies, informed by LCA evidence
- Policy monitoring of relevant health indicators to mitigate unintended harms and to iteratively inform research

## Materials and Methods

Our review is conducted according to the standardised technique for assessing and reporting reviews of life cycle assessments (STARR-LCA) checklist (*86*), based closely on the Preferred Reporting Items for Systematic Reviews and Meta-Analyses (PRISMA) reporting guidelines(*87*). A PRISMA checklist is available in **Table S1**. A study protocol was registered with Prospero international prospective register of systematic reviews (*88*).

### Search strategy

We systematically searched Web of Science, Scopus, Medline, EMBASE, Global Health and GreenFile databases using key search terms for life cycle assessments and packaging types, combined with more specific terminology to identify assessments of food and drink packaging, particular plastic materials and recycling and reuse comparisons. Database searches were run on 29^th^ July 2020 and updated on 22^nd^ July 2021 with no limits on geographical location, language or publication date. An example search string for Scopus is provided in full and was adapted only where necessary for the functionality of other databases. All search strings are provided in **Tables S2-S7**.

*TITLE-ABS-KEY ((lca OR lcas OR lcia OR lcias OR {life cycle} OR {life-cycle}) AND (packag* OR pack OR packs OR packet OR packets OR container* OR casing OR casings OR pouch OR pouches OR pot OR pots OR punnet OR punnets OR carton OR cartons OR box OR boxes OR tray* OR film OR films OR wrap OR wraps OR wrapping OR wrappings OR wrapper OR wrappers OR bottle* OR cup OR cups OR bag OR bags OR cap OR caps OR clamshell* OR tableware OR {food service ware} OR cutlery OR crockery OR spoon OR spoons OR fork OR forks OR knife OR knives) AND (food* OR beverage* OR drink* OR water OR waste OR plastic OR plastics OR polyolefin OR polyolefins OR polyethylene OR polythene OR pe OR pet OR hdpe OR mdpe OR ldpe OR lldpe OR {polyvinyl chloride} OR pvc OR polypropylene OR pp OR polystyrene OR ps OR acrylic OR polycarbonate OR pc OR polylactide OR {polylactic acid} OR pla OR styrofoam OR styrene OR {acrylonitrile butadiene} OR nylon OR pa OR fibreglass OR fiberglass OR tetrapak OR {tetra pak} OR {single-use} OR {single use} OR {one-way} OR {one way} OR disposable OR vacuum OR aseptic OR multilayer* OR recycl* OR reuse OR reusing OR reused OR reusable OR multi-use OR multiuse))*

Websites of relevant organisations and select government agencies were searched for grey literature including: the United Nations Environment Programme Life Cycle Initiative, the European Platform on Life Cycle Assessment, the European Environment Agency, Waste and Resources Action Programme (WRAP), the United Kingdom government website, EThOs database for doctoral research and a Google search for additional government agency studies. Reference lists of included LCAs were searched and authors were contacted where necessary to retrieve additional or related studies.

### Eligibility criteria

We included peer-reviewed, process-based, comparative life cycle impact assessments of plastic food and drink primary packaging, food service ware and grocery bags that compare virgin and single use plastics with recycling and reuse options. Eligible functional units included those based on a specific number of packaging items (e.g. 1000 cups), a measure of packaging weight (e.g. 1000kg of plastic food packaging) or the provision of a service (e.g. the bags required for 1000 shopping trips).

The Resin Identification Codes for the most common plastics were used to inform the search string: Polyethylene Terephthalate, High-Density Polyethylene, Polyvinyl chloride, Low-Density Polyethylene, Polypropylene and Polystyrene, Polycarbonate, Acrylic, Fiberglass, Nylon, Acrylonitrile Styrene and Polylactic Acid (*89*). Other types of plastic, including bio-based plastics were included where identified in the literature. Primary packaging was defined as consumer-level food or drink contact packaging, service ware included any items such as crockery or cutlery for takeaway food and drink consumption and bags were included if specified for groceries. These items were collectively referred to as ‘packaging’ in this review.

Including only comparative LCAs, focusing on internal differences in impacts, reduces confounding by varying methodological choices and assumptions between studies(*86*).

Three groups of internal comparisons were eligible:

1. Recycled content of plastic packaging: LCAs that compare a unit of packaging made entirely of virgin plastic, or lower recycled content, compared to the same made of any higher level of recycled plastic content. For example, a virgin polyethylene terephthalate (PET) bottle compared with a 20% recycled content PET bottle
2. Recycling as an end-of-life plastic packaging scenario: LCAs that compare incineration and/or landfill of plastic packaging at the end of its lifespan with any higher rates of recycling. For example, a PET bottle with 100% incineration at the end of its lifespan compared to 100% recycling
3. Reusable plastic packaging: LCAs that compare plastic packaging designed for single use compared to packaging designed to be reused and for the same purpose. These studies may compare any number of uses of the reusable product. For example, a PET bottle designed for single use, and used once, compared with a PET bottle that is designed to be reused, and is used 20 times

Eligible studies considered the whole packaging life cycle, at a minimum including both plastic production and end-of-life stages in the scope of the study.

LCA methodology offers a range of impact categories, from which individual studies may select any number to model. To be eligible for our review, each LCA was required to model at least one of the available health-related impact categories including: the endpoint area of protection category “Human health” (calculated in Disability-Adjusted Life Years) and/or the predefined contributing “Human health” midpoint impact categories including “Climate change”, “Stratospheric ozone depletion”, “Particulate matter formation”, “Photochemical ozone formation”, “Ionizing radiation”, “Human toxicity” and “Water Use” (*18*). This terminology is taken from the ReCiPe 2016 LCA impact assessment method (*90*); the specific health effects encapsulated by each category are detailed in **Figure 1** and relate mainly to morbidity and mortality from respiratory diseases and cancers, and malnutrition, malaria, diarrhoea, and flooding. Various terminologies for these impact categories exist in different LCA impact assessment methods; all were eligible for our review and are conceptually comparable to those listed.

Studies that only reported life-cycle inventory data, purely economic input-output or social LCAs and all other study designs were excluded. LCAs that only compared plastic packaging with another type of material (e.g., glass or paper) and those in which the packaging was a composite of plastic and other materials such as paperboard were excluded. Packaging used in other parts of the food system (e.g., in agriculture or secondary and tertiary packaging for bulk transport) was excluded, as were studies in which packaging was only part of the inventory list for a food product functional unit. Studies that only reported environmental, non-health related impact indicators according to LCA methods were excluded, for example Resource Depletion and Eutrophication(*18*).

### Data screening and extraction

Records were exported from database searches into Eppi Reviewer software. Screening was conducted in two stages by MD, first on title and abstract and then by full text, with 10% checks completed by HR and JY at both stages. Agreement rates were 98% at title and abstract stage and 94% on full text, discrepancies were discussed and resolved by consensus.

The data extraction form was developed in Eppi Reviewer to capture publication date and geographical focus, compliance with the International Organization for Standardization (ISO) or other LCA standards, the LCA goal and scope, functional unit, specific packaging function and material composition, the system boundaries, specific recycling and/or reuse comparison, the life cycle inventory data sources, key assumptions, recycling and reuse allocation methods, impact assessment software and methods, impact categories and indicators and the absolute impact values or relative differences from study comparisons. Data extraction was carried out by MD with 10% checks completed by XY. Study authors were contacted directly to request additional information and data where necessary.

### Reporting Quality and Suitability for Meta-analysis

We only included LCAs that were peer-reviewed or subject to critical review, as specified in the ISO LCA standards for comparative life cycle assessments (*91, 92*), to ensure quality evidence with the lowest risk of bias. In addition to this selection criterion, we adapted a tool proposed by Price and Kendall (*93*) to assess the reporting transparency of included articles and their suitability for the purpose of our meta-analysis. This is not an appraisal of overall study quality, and it should be noted that lower scores may indicate that individual studies, whilst relevant, are designed to meet different objectives from those of our review. This assessment was carried out on all studies by MD with 10% checks completed by XY. The full tool is provided in **Figure S6**.

This tool considers three domains of reporting quality and suitability for meta-analysis:

1. Completeness of life-cycle modelling: LCAs were graded on a ten-point scale according to their consideration of the complete life cycle of packaging, including raw material extraction, product manufacturing, transport and distribution, consumer use and end-of-life scenarios, and whether evidence of use of sufficient data was provided for each of these stages
2. Methodological focus and transparency: Focus was defined as the alignment of individual LCA objectives with those of our review, in terms of assessing health outcomes. Transparency was graded on a three-point scale, based on the provision of a clear description of the functional unit with specific material weights, the method of accounting and crediting systems for recycling and reuse, and the impact assessment methodology used
3. The granularity of presented results: LCAs were graded on a six-point scale based on their reporting of overall life cycle impacts, and whether results were disaggregated by life cycle stages

Given the limited number of studies, the resulting scores were not used to exclude studies from meta-analysis but to give an overall indication of quality and to aid discussion of discrepancies in results.

### Analysis

#### 1) Evidence Map

Using Eppi Reviewer Software, an interactive evidence and gap map was created to visualise the frequencies of LCAs according to the three circular economy comparison domains: 1) Recycled content, 2) End-of-life recycling and 3) Reuse. This was cross tabulated with the frequencies of LCA impact categories modelled in the literature. The map is segmented by product type and can be further filtered by plastic material types. A static version of this map was created using Excel.

#### 2) Average percent change in impacts for recycled content, recycling, and reusable packaging

Results from individual studies were tabulated and analysed in Excel. Where studies reported multiple relevant comparisons, all were extracted and used as individual comparisons in the analysis. These are referred to as ‘comparison points’ throughout our review. For recycled content and end-of-life scenarios, comparisons were restricted to those looking at the same type of plastic material to avoid confounding of the effects by different resource requirements and emissions related to different types of plastic. For reuse comparisons however, different types of material comparisons were permitted to account for the predominantly different design, often in terms of material type and weight, of reusable products.

For each impact category assessed, we tabulated the absolute value for the reference product or scenario, the absolute value for the comparison product or scenario, the absolute difference and the relative percent change between the two. Where studies reported only the absolute values for each of the products or scenarios, we calculated the absolute difference and relative percent change using the following formulas:

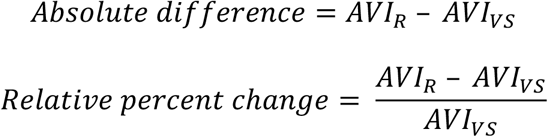

Where:

*AVI*_*VS*_ = The absolute impact category value for the reference virgin plastic or single use plastic product or scenario

*AVI*_*R*_ = The absolute impact category value for the recycling or reuse comparison

Some LCAs result in negative impacts, this is where environmental credits awarded to the system outweigh environmental burdens and the product is perceived to provide a net-benefit. For example, if the burdens of packaging production are less than the energy recovery from incineration credits awarded for electricity generation. Percent change comparison points where one of the starting values was negative were excluded from this meta-analysis because these values were not meaningful (*94*). Where absolute values were both negative, the relative percent change was included. Excluded calculations are provided in the Supplementary Data File and highlighted in red.

Impact categories resulting from different LCA impact assessment methods were grouped based on similarity. For consistency, we retain the terminology used in ReCiPe 2016 impact assessments though the resulting groups may contain indicators such as the following:

- **Climate Change (kg CO**_**2**_ **equivalents):** Global Warming Potential (kg CO_2_ equivalents), Carbon Footprint (kg CO_2_)
- **Stratospheric Ozone Depletion (kg CFC-11 equivalents):** Ozone Depletion Potential (kg R11 equivalents)
- **Photochemical Ozone Formation (kg NO**_**x**_ **equivalents):** Ozone Creation Potential, Photochemical Oxidant Formation (kg NMVOC equivalents), Respiratory Organics (kg C2H4 equivalents, Summer Smog (kg ethene equivalents)
- **Ionizing Radiation (kBq Co-60 to air eq/kBq):** Ionizing Radiation (kBq U235-equivalents)
- **Fine Particulate Matter Formation (kg PM2.5 equivalents):** Respiratory Inorganics (kg PM2.5 equivalents)
- **Human Toxicity (ReCiPe 2008) (kg 1**,**4-DCB equivalents):** Human Toxicity Potential (kg DCB equivalents)
- **Human Toxicity Cancer Effects (ReCiPe 2016) (kg 1**,**4-DCB equivalents):** Human Carcinogenic Toxicity (kg 1,4 DCB), Carcinogens (kg C2H3Cl equivalents), Carcinogenic risk (mg As equivalents)
- **Human Toxicity Non-Cancer Effects (ReCiPe 2016) (kg 1**,**4-DCB equivalents):** Non-carcinogens (kg C2H3Cl equivalents)
- **Water Use (m**^**3**^): Resource Depletion Water (m^3^ equivalents), Water Consumption (L), Water Depletion (gal), Water Scarcity Indicator (m^3^)

Relative measures are useful for their robustness to differences between studies in terms of the functional unit and specific impact indicators. They also reflect the greater strength of LCAs in comparing products and scenarios rather than accurately assessing absolute emissions(*91, 92*). For this reason, we first summarised the relative percent change within restricted scenarios that were most frequently assessed in the literature:

1. Recycled content: Virgin plastic (0% recycled content) vs 100% recycled content
2. End-of-life recycling:
  a. 100% landfill vs 100% recycling
  b. 100% incineration vs 100% recycling
3. Reuse: Products designed for reuse vs products designed for single use based on a one-time use

The impacts of these scenarios were summarised using the median and interquartile range of the relative percent change values calculated from individual comparisons within studies. This reflects the average relative change in specific emission categories across food packaging types under scenarios of switching entirely from virgin plastic to recycled plastic, from landfill and incineration to complete recycling and from single-use design to reusable design based on one use. The mean and standard deviations are available in **Figures S7-S10**.

### Meta-analysis using mixed-effects linear regression

We used mixed-effects linear regression, specifying individual studies as the random effects to account for multiple comparison points from each study, and using the range of values found in the literature for percentage recycled content, end-of-life recycling rates and number of uses for reusable products as predictor variables.

Including only studies that used 0% recycled content, 0% recycling or a single use item used once respectively as the reference baseline allowed the response variable to be meaningful as the absolute or relative difference in LCA health-related impacts. In line with Cochrane advice, we also limited meta-regressions to LCA impact categories for which there was minimum of 10 studies (*95*), and that had at least 5 variations in the predictor variable. For example, at least 10 studies that considered Climate Change impacts of increasing recycled content of plastic packaging, relative to virgin packaging, with at least 5 variations in the percentage of recycled content across available comparison points.

For both recycled content comparisons and end-of-life recycling comparisons, we were able to calculate meaningful absolute differences in impacts because internal study comparisons were restricted to those considering the same type of plastic with the same weight for the virgin plastic product and the recycled content product. To make the absolute differences comparable across studies we standardised functional units to 1 metric tonne of plastic packaging using the material weights provided by each study.

All midpoint impacts were standardised to ReCiPe 2016 Hierarchic perspective midpoint indicators, and subsequently converted to DALYs using the same method (*90*). The only exceptions were for some midpoint Human Toxicity indicators for which 2016 conversions were not available. We converted Comparative Toxic Units for human toxicity (CTUh) to DALYs using factors for Cancer Effects and Non-Cancer Effects from the International Resource Panel (*96*). Where “Human Toxicity” was not disaggregated by Cancer and Non-Cancer Effects, it was necessary to use the ReCiPe 2008 conversion factor(*97*). This is likely to underestimate DALYs and was highlighted in the limitations. Where converting impact indicators was not possible or meaningful, these data were excluded from the meta-analysis, these data are highlighted in the Supplementary File.

Studies reported different numbers of health-related midpoint indicators. The Total DALYs is based on the sum of DALYs from all provided midpoint indicators in each study. This means that for some studies, DALY estimates are based on one pathway only e.g. health effects of Climate Change, whereas estimates from other studies will be based on multiple pathways e.g. health effects of Climate Change, Particulate Matter Formation and Human Toxicity combined, depending on what data was provided in the study. A detailed description of all conversions and the number of midpoint indicators contributing to each DALY estimate is provided in the Supplementary file.

For comparisons of reuse, only the relative precent change was used in regressions because the differences in material composition and weight between single-use and reusable products within studies, and the different definitions of functional units between studies meant that standardising to calculate the absolute difference was not possible. Instead, the number of uses required to ensure the reusable product had lower or equal health related impacts compared to the single use product was estimated using the regression formula to calculate x (number of uses) when y=0 (relative percent change in impacts). This is referred to as the ‘break-even point’.

All regressions were run in Stata 17.0 using the *xtmixed* command with random effects for study name. Regressions for recycled content and end-of-life recycling included a fixed intercept at zero. This is because the only explanatory variable was percentage recycled content or percentage end-of-life recycling and the response variable was the absolute difference in impacts with a fixed reference point of 0% recycled content and 0% recycling respectively. Therefore, given that comparison points were based on the same products, with the same weight and material type, when x=0 for these regressions, the absolute difference (y) must also equal zero and fixing the intercept permits the coefficient for the change in absolute difference to be meaningfully interpreted. Random effects from multiple comparison points in individual studies were applied to the slope. This was not done for the reuse model because the same inclusion criteria for comparison points of equal material type and weight do not apply in this analysis. The Stata do file is available in **Data S12** and all Stata regression outputs are available in **Figures S11-S13**.

## Data Availability

All data produced in the present study are available upon reasonable request to the authors

## Acknowledgments

We would like to thank R. Burke at LSHTM for advising on the search strategy, E. Hutchinson from LSHTM for consulting on the analysis of health outcomes and M. Huijbregts for advising on ReCiPe 2016 conversion factors and the analysis strategy. With thanks also to M. E. Correa Cano at Exeter University, and A. Gasparrini and D. Macleod at LSHTM for advice on statistical queries. We would like to acknowledge the EPPI Reviewer team at the University College London, which includes J.Brunton, Z. Grouze, S. Graziosi and M. Bond.

## Author contributions

Conceptualization: MD, JY, SK, RG, XY

Methodology: RG, XY, MD, CD, SK

Investigation: MD, XY, CD, JY, HR, RG

Visualization: MD, RG, CD

Funding acquisition: SK, JY

Project administration: SK, JY

Supervision: SK, RG

Writing – original draft: MD

Writing – review & editing: MD, SK, RG, CD, JY, HR

## Data and materials availability

All data are available in the main text or the supplementary materials.

## Supplementary Materials

**Supplementary Materials File**

Figs. S1 to S13

Tables S1 to S7

References (1 to 1)

**Other Supplementary Materials for this manuscript include the following:**

**Data S1-S11** (separate file)

**Data S12** (separate file)

